# Irritancy and spatial repellency efficacy of repellent-treated fabrics against *Aedes aegypti* (L.) (Diptera: Culicidae) in an excito-repellency system

**DOI:** 10.1101/2025.10.31.25339223

**Authors:** Alex Ahebwa, Jeffrey Hii, Theerachart Leepasert, Jirod Nararak, Monthathip Kongmee, Theeraphap Chareonviriyaphap

## Abstract

Mosquito-borne diseases remain a major public health challenge, driving the need for affordable and scalable vector control tools. In this study, we used an excito-repellency system to evaluate the deterrent potential of two low-cost repellent-treated fabrics consisting of Calico (100% cotton) and Jute (hessian), to a standard treated bed net polyester (BNP), against a laboratory strain of *Aedes aegypti* (L.). Fabrics (15 x 17.5 cm) were treated with six concentrations of transfluthrin and metofluthrin, and five of permethrin, categorized as low and high doses. Chemical retention was assessed using gas chromatography–mass spectrometry (GC-MS), and behavioral avoidance was measured via chamber escape responses over 30 minutes using 60 female mosquitoes per replicate (n = 4). Calico elicited the strongest mosquito escape (OR = 3.12) followed by Jute (OR = 1.74). Transfluthrin (OR = 5.45) produced the most escape whereas low dose treatments resulted in greater escape (OR = 1.23) than high dose applications. Non-contact chambers elicited more escape (OR = 1.87) than the contact test chamber, indicating stronger spatial repellency than irritancy. GC-MS results confirmed fabric-specific chemical retention dynamics: permethrin remained stable across all fabrics and time points, while transfluthrin retention varied significantly between Calico and BNP after 24 h drying. These findings highlight the importance of fabric– insecticide compatibility and the influence of exposure method, dose, and chemical volatility on repellent efficacy. Notably, both metofluthrin-treated fabrics and BNP exhibited pronounced insecticidal effects, suggesting that toxicity may mask repellency—a hypothesis warranting further evaluation at lower concentrations.

## Introduction

*Aedes aegypti* is the most widely distributed mosquito vector globally, responsible for transmitting arboviruses of major public health concern, including dengue, chikungunya, Zika, and yellow fever [1, 2]. The rising incidence of these diseases is driven by climate change, urbanization, global travel, and expanding trade networks [3, 4]. In the absence of widely accessible vaccines and reliable diagnostics, mosquito bite prevention and vector control remain the cornerstone of disease mitigation [5].

While insecticide-treated nets (ITNs) and clothing (ITC) are effective against malaria vectors [6, 7], their efficacy against diurnal vectors like *Ae. aegypti* is inconsistent [8]. Given *Ae. aegypti*’s daytime activity, when ITN use is minimal, ITC offers a practical alternative, especially in occupational and recreational settings [9, 10]. Their integration public health settings may be facilitated by occupational norms, such as protective clothing worn by rubber tappers [11] and the availability of guidelines for home-treatment [12]. Recent studies have explored a range of textile substrates, including natural fibers like cotton and jute, and synthetic blends such as polyester and nylon, each with distinct insecticide retention and repellent profiles [13–17]. Natural fibers such as cotton and jute have also been evaluated for their compatibility with spatial repellents, offering low-cost alternatives with variable chemical retention [12, 14, 16, 18]. Innovations in mosquito-repellent textiles now extend to outdoor wear, uniforms, and bedding, with efficacy shaped by fabric porosity, weave density, and treatment method [12]. Despite these advances, standardized guidelines for fabric selection and treatment remain limited.

Permethrin is the primary insecticide used in ITCs and acts through contact-mediated irritancy, disrupting mosquito landing behaviour [19, 20]. When combined with a topical repellent, near-complete protection has been achieved [21–23]. Still, exposed skin remains vulnerable, particularly in regions with insecticide-resistant vector populations. Volatile pyrethroid spatial repellents (VPSRs) such as transfluthrin and metofluthrin, offer an added layer of protection with olfactory-mediated repellency that bypasses traditional resistance mechanisms [5, 24, 25]. Despite this promise, empirical validation of VPSR-impregnated fabrics remains limited. It is reasonably difficult to measure repellency under field conditions or even semifield conditions given the nature of the VPSRs [26–28]. The excito-repellency system (ERS)—which distinguishes spatial repellency (non-contact chamber) from contact irritancy (contact chamber)—provides a robust framework for evaluating mosquito behavior under laboratory conditions [29, 30].

This study, therefore, used ERS to evaluate the irritancy and spatial repellency of two low-cost fabrics—calico 100% calico and jute [hessian/burlap]—impregnated with either transfluthrin, metofluthrin or permethrin. Their performance was compared to standard bed net polyester. The retention of transfluthrin and permethrin in the fabrics before and after the drying period was quantified using the gas chromatography – mass spectrometry (GC-MS).

## Materials and methods

### Mosquito rearing

*Aedes aegypti* (USDA), a laboratory-adapted susceptible strain that has been maintained for over 20 years at Kasetsart University, Bangkok, Thailand was used for all bioassays. Rearing followed Ahebwa et al [14]: 26 ± 2 °C, 70 ± 10% relative humidity, and a 12:12 h light:dark cycle. Eggs were hatched in tap water (2 L/tray), and larvae were reared at a density of 250 per tray, fed daily on commercial food pellets (PondMax, Australia; 2 pellets ≈100–150 mg per tray). Pupae were transferred to 30 × 30 × 30 cm cages for adult emergence. Adult were sustained on 10% sugar solution through calico swabs; females were blood-fed twice weekly using CPDA-1 preserved human blood (from Thai Red Cross Society) via artificial membrane feeding. Moistened filter papers were introduced into each cage to encourage oviposition, and dried eggs were stored for colony maintenance. Unfed, 3–5 day old female adults were selected for assays after 24 h sugar withdrawal.

### Volatile pyrethroid preparation and dose-ranging

Technical-grade transfluthrin (97.9%; CAS 118712-89-3), metofluthrin (96.4% purity; CAS 240494-70-6) (Earth (Thailand) Co. Ltd., Bangkok), and permethrin (94%) were used to prepare stock solutions. Analytical-grade acetone (Avantor Performance Materials, Inc., Allentown, PA, USA) was used as an organic solvent, and silicone oil (Dow Corning1556, Dow Chemical Co., Midland, MI, USA) as a carrier at a 0.96:1.95 ratio [14]. Preliminary dose-range optimization was conducted using calico fabric swatches (15 x 17.5 cm) impregnated with 3 mL insecticide solution. Reference discriminating concentrtations were 0.06824% w/v for transfluthrin [31] and 0.4% w/v for permethrin [32]. Subsequently, six transfluthrin (and metofluthrin) concentrations (1.0–31.3 mg/m^2^) and five for permethrin (0.25–4 g/m^2^) were prepared by serial dilution.

### Fabric preparation and treatment

Three fabric types were sourced from private suppliers in Bangkok, Thailand comprising Calico (100% cotton), Jute (100% jute yarn fabric), and BNP (bed net polyester). Prior to use, all fabrics were screened for insecticide contamination using WHO cone bioassays with 200 laboratory-susceptible *Ae. aegypti* females. Any fabric yielding >5% mortality 24 h was excluded. Each fabric was cut into eight 15 x 17.5 cm pieces: four for contact and four non-contact chambers in the excito-repellency system. Four untreated control pieces were reused across bioassays. Fabrics were treated with 3 mL of insecticide solution using calibrated 10 mL glass pipettes, following WHO guidelines [33]. Control pieces received an equivalent volume of solvent mixture. Treated fabrics were air-dried for 24 h at 26 ± 4 °C, 70 ± 10% relative humidity under a 12:12 light:dark photoperiod. Treated fabrics were discarded after each test.

### Excito-repellency bioassays

The improved excito-repellency system, based on the original design by Chareonviriyaphap et al [34], with modifications described in our recent studies [35, 36], distinguishes between contact irritancy and non-contact spatial repellency **(Fig 1)**. In this study, treated fabrics replaced the standard Whatman No. 1 filter papers. Each bioassay consisted of a single fabric type and insecticide concentration tested against 60 adult female *Ae. aegypti* in four replicates of 15 mosquitoes. After a 3-min acclimation period, exit doors were opened into adjacent receiving boxes. Escaping mosquitoes were recorded over 30 min. Escaped and non-escaped mosquitoes were transferred to labeled, net-covered paper cups and provided 10% sugar solution. Mortality was recorded 24 h later. Room conditions were maintained at 25 ± 2 °C and 70 ± 5% relative humidity throughout the experiment. Chambers were cleaned with acetone between tests and fully submerged in acetone overnight between insecticides, then air-dried for 24 h before reuse.

**Fig 1.**
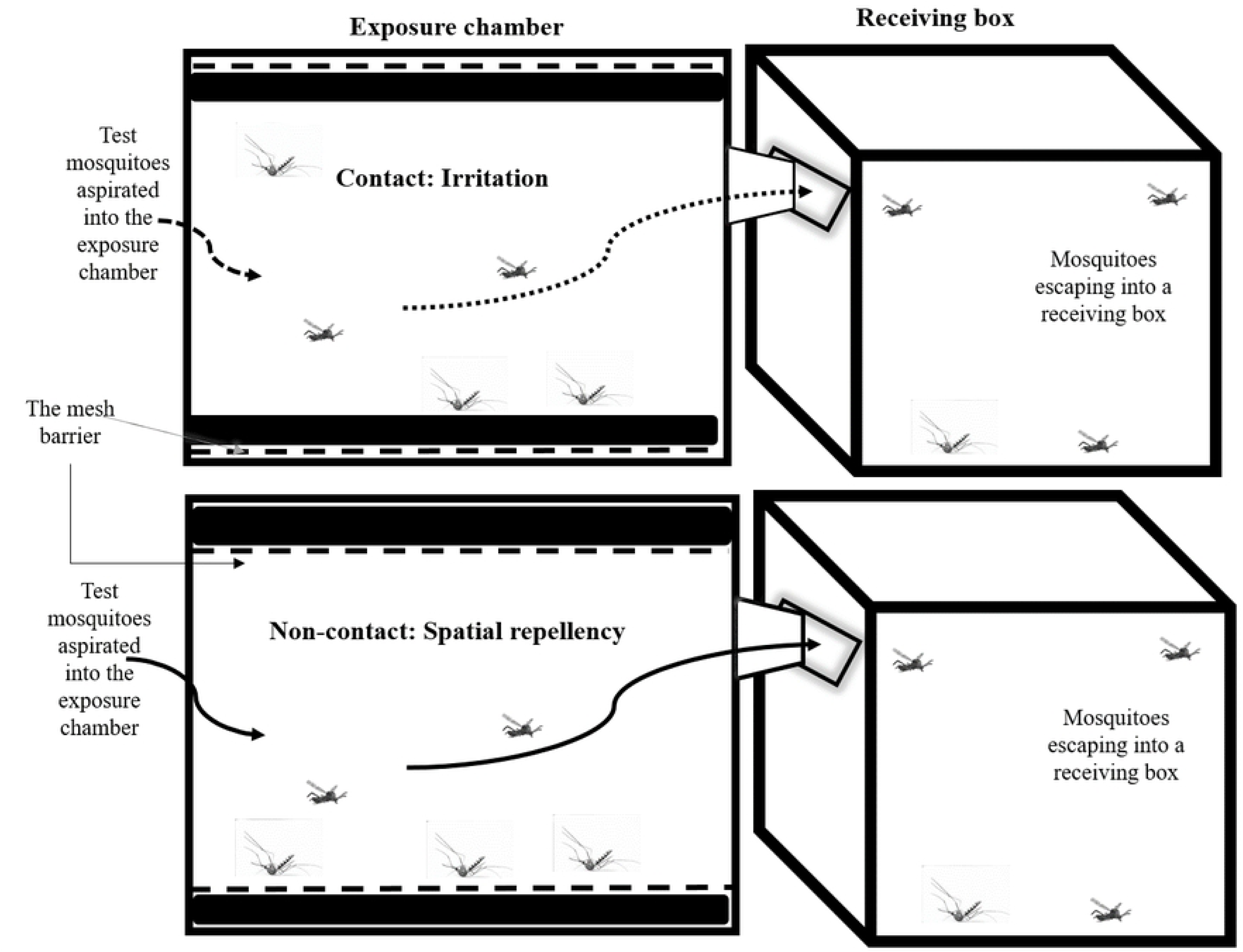
Cross-sectional view of the excito-repellency system showing the contact exposure chamber (upper left) and non-contact exposure chamber (lower left). Each chamber is connected to a receiving box, which collects mosquitoes that escape due to irritation or spatial repellency. Mosquitoes remaining in the exposure chambers are recovered after each bioassay, while those that escape are collected immediately upon exit.

### Chemical analysis

Gas chromatography–mass spectrometry (GC-MS) was employed to assess transfluthrin and permethrin retention in Calico, Jute, and BNP fabrics after 1 h and 24 h drying intervals, under controlled laboratory conditions (25 ± 2 °C; 70 ± 10% RH). Previous findings suggest that transfluthrin concentrations decline over time, potentially reducing efficacy [37]. Following impregnation, each insecticide was applied to a single 15 × 17.5 cm fabric piece. Three 4 cm² swatches were excised at each time point for chemical extraction. Swatches were soaked overnight in analytical-grade methanol and sonicated for 10 min; extracts were transferred to glass vials for GC-MS analysis.

A QP2020 gas chromatography system (Shimadzu, Japan) with SH-Rxi-5Sil MS column (30 m × 0.25 mm i.d., 0.25 µm film) was used. Helium was used as the carrier gas at 1.2 mL/min with a split ratio of 1:10. The injector was maintained at 230 °C. Oven temperature was programmed from 50 °C (initial) to 220 °C at a ramp rate of 4 °C/min. Selected ion monitoring (SIM) was employed for compound detection. Target ions were m/z 163 and 91 for transfluthrin [38, 39], and m/z 183, 163, and 165 for both cis- and trans-permethrin [40].

### Data analysis

Raw data were entered in Microsoft Office Excel (Windows 11) and imported into RStudio version 4.1.0 [41] for statistical analysis. Data manipulation and cleaning were accomplished using the *tidyr* package [42]. The escape rates were adjusted with the paired controls to set the controls at zero based on the Abbott’s formula [43].

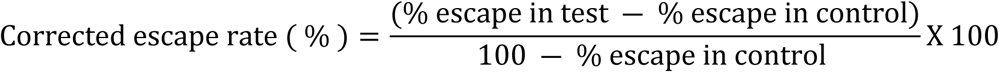

To assess the escape patterns, Kaplan-Meier survival curves were generated using the `survfit` function from the `survival` package, providing non-parametric estimates of survival probabilities (number of mosquitoes that did not escape) over time, with escape as the event. Visualizations were produced with `ggsurvplot` from the `survminer` package. Differences among groups were evaluated using the log-rank test, with statistical significance defined as *p* < 0.001. Right-censored data (i.e., mosquitoes that did not escape within the observation period) were appropriately accounted for [44].

To model the probability of mosquito escape as a function of fabric type, insecticide type, and exposure mode (contact vs non-contact), a generalized linear mixed model (GLMM) was fitted using the `glmer` function from the `lme4` package. The response variable—number of mosquitoes released per replicate—was binary (escaped vs not escaped), with fixed effects for fabric type, exposure mode, and insecticide type, and random effects for insecticide dose and replicate trials. Model fit was evaluated using Akaike Information Criterion (AIC), and statistical significance of predictors was assessed via likelihood ratio tests and Wald statistics [45]. Model outputs— including estimated coefficients, odds ratios (ORs), 95% confidence intervals (CIs), and P-values—were summarized in tabular format to facilitate interpretation of fixed effects and their relative influence on escape behavior. Odds ratios and 95% confidence intervals were derived by exponentiation of model coefficients.

For analysis purposes that focuses on the efficacy of the treated fabrics, insecticide doses were categorized as low (first three doses) or high (remaining doses) based on treatment concentration ranges. This dose classification allowed for consistent comparison across insecticides and fabrics, while retaining operational relevance for field application.

## Results

### Irritancy and spatial repellency

Mosquito escape behavior was evaluated using generalized linear mixed models (GLMM) and Kaplan-Meier survival analysis, with escape treated as the event. Irritancy was defined by escape in contact trials, while spatial repellency was inferred from escape in non-contact trials.

In the GLMM, reference levels were set as metofluthrin (insecticide), BNP (fabric), high dose, and contact chamber (Intercept: OR = 0.052; 95% CI: 0.039–0.070). All main effects significantly influenced escape behavior (*P* < 0.001), except for insecticide–fabric interaction terms. Relative to BNP, Calico significantly increased escape odds (OR = 3.12; 95% CI: 2.24– 4.34), followed by Jute (OR = 1.74; 95% CI: 1.22–2.47). Transfluthrin elicited the strongest escape response (OR = 5.45; 95% CI: 3.96–7.50), with Permethrin also showing elevated escape (OR = 3.26; 95% CI: 2.32–4.58) compared to metofluthrin. Notably, the transfluthrin–Calico interaction reduced escape odds by 49% (OR = 0.51; 95% CI: 0.35–0.77), while the permethrin–Calico interaction increased escape by 45% (OR = 1.45; 95% CI: 0.95–2.20). Escape was significantly higher in non-contact trials (OR = 1.87; 95% CI: 1.65–2.11) and at low dose levels (OR = 1.23; 95% CI: 1.09–1.38), suggesting dose-dependent spatial repellency (Table 1).

**Table 1.**
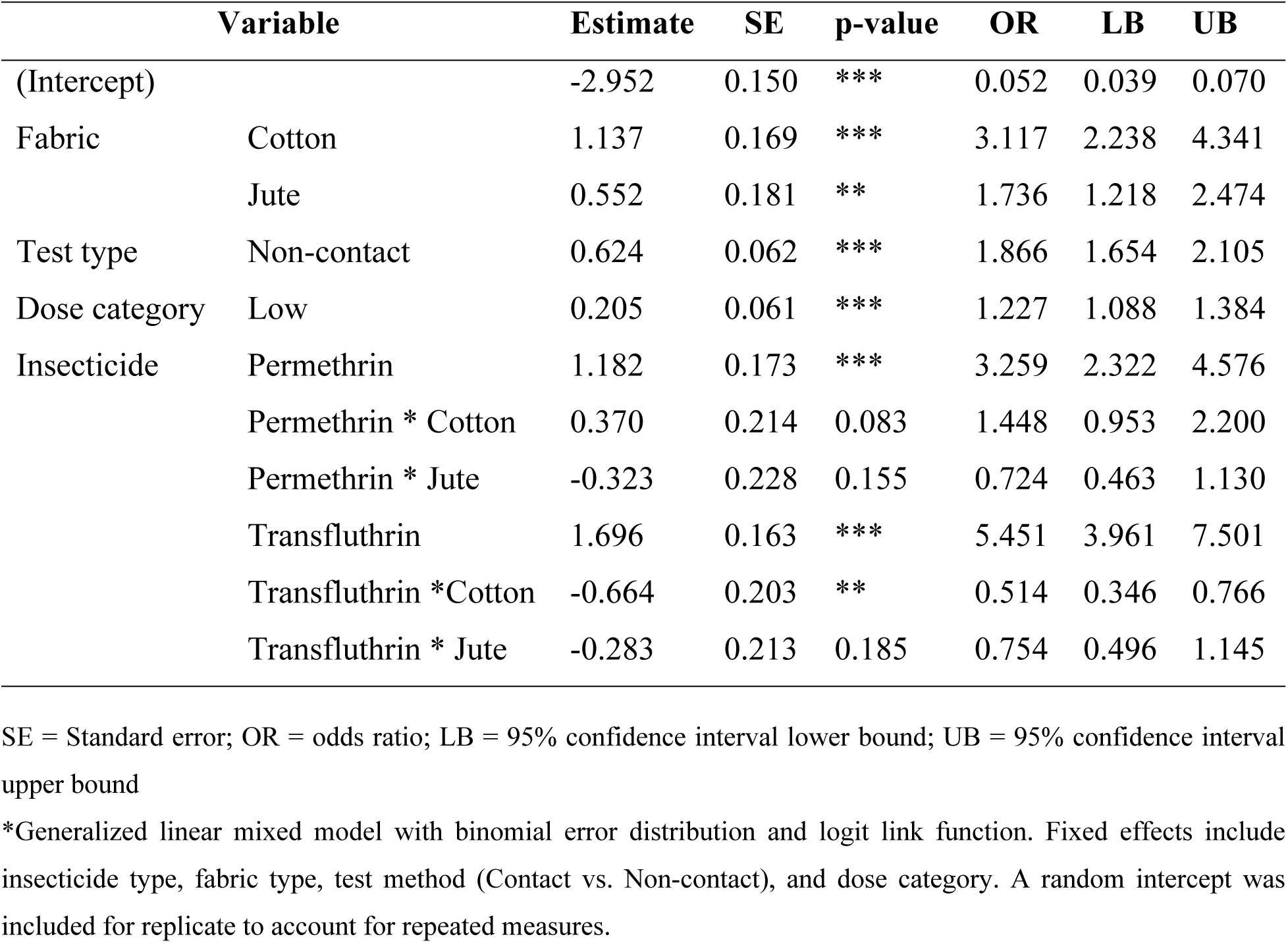
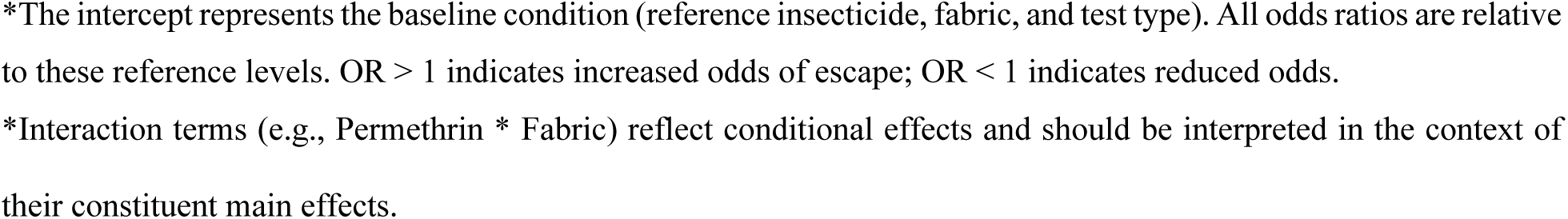
Estimated effects of insecticide, fabric type, and test method on *Aedes aegypti* escape behavior using binomial GLMM.

| Variable |  | Estimate | SE | p-value | OR | LB | UB |
| --- | --- | --- | --- | --- | --- | --- | --- |
| (Intercept) |  | -2.952 | 0.150 | *** | 0.052 | 0.039 | 0.070 |
| Fabric | Cotton | 1.137 | 0.169 | *** | 3.117 | 2.238 | 4.341 |
|  | Jute | 0.552 | 0.181 | ** | 1.736 | 1.218 | 2.474 |
| Test type | Non-contact | 0.624 | 0.062 | *** | 1.866 | 1.654 | 2.105 |
| Dose category | Low | 0.205 | 0.061 | *** | 1.227 | 1.088 | 1.384 |
| Insecticide | Permethrin | 1.182 | 0.173 | *** | 3.259 | 2.322 | 4.576 |
|  | Permethrin * Cotton | 0.370 | 0.214 | 0.083 | 1.448 | 0.953 | 2.200 |
|  | Permethrin * Jute | -0.323 | 0.228 | 0.155 | 0.724 | 0.463 | 1.130 |
|  | Transfluthrin | 1.696 | 0.163 | *** | 5.451 | 3.961 | 7.501 |
|  | Transfluthrin * Cotton | -0.664 | 0.203 | ** | 0.514 | 0.346 | 0.766 |
|  | Transfluthrin * Jute | -0.283 | 0.213 | 0.185 | 0.754 | 0.496 | 1.145 |

SE = Standard error; OR = odds ratio; LB = 95% confidence interval lower bound; UB = 95% confidence interval upper bound
\*Generalized linear mixed model with binomial error distribution and logit link function. Fixed effects include insecticide type, fabric type, test method (Contact vs. Non-contact), and dose category. A random intercept was included for replicate to account for repeated measures.
\*The intercept represents the baseline condition (reference insecticide, fabric, and test type). All odds ratios are relative to these reference levels. OR > 1 indicates increased odds of escape; OR < 1 indicates reduced odds.
\*Interaction terms (e.g., Permethrin \* Fabric) reflect conditional effects and should be interpreted in the context of their constituent main effects.

Kaplan-Meier survival curves supported these findings. Metofluthrin produced early plateauing curves in contact trials at both low and high dose levels (survival: >85% and 100%, respectively), indicating limited irritancy (Fig S1). In non-contact trials at low dose, survival declined gradually, with Calico showing the steepest initial drop and reaching 42.2% survival (57.8% escaped) (Fig 2). Permethrin-treated Calico consistently exhibited strong irritancy and spatial repellency. In contact trials, survival dropped to 50% within 10 minutes at both dose levels, and further declined to 31.7% at the high dose, indicating the strongest overall irritancy (68.3% escaped; Fig 3). In non-contact trials at the high dose, survival decreased to 30.8%, reflecting the strongest overall spatial repellency (69.2% escaped; Fig 4). With transfluthrin, fabrics showed no significant differences in mosquito survival (P = 0.16), with overlapping survival curves across Jute and Calico (Fig 5). In contrast, non-contact trials at the high dose showed peak spatial repellency, with survival dropping to 75% within 5 minutes for both fabrics and further declining to 40% and 36.1%, respectively (Fig 6).

**Fig 2.**
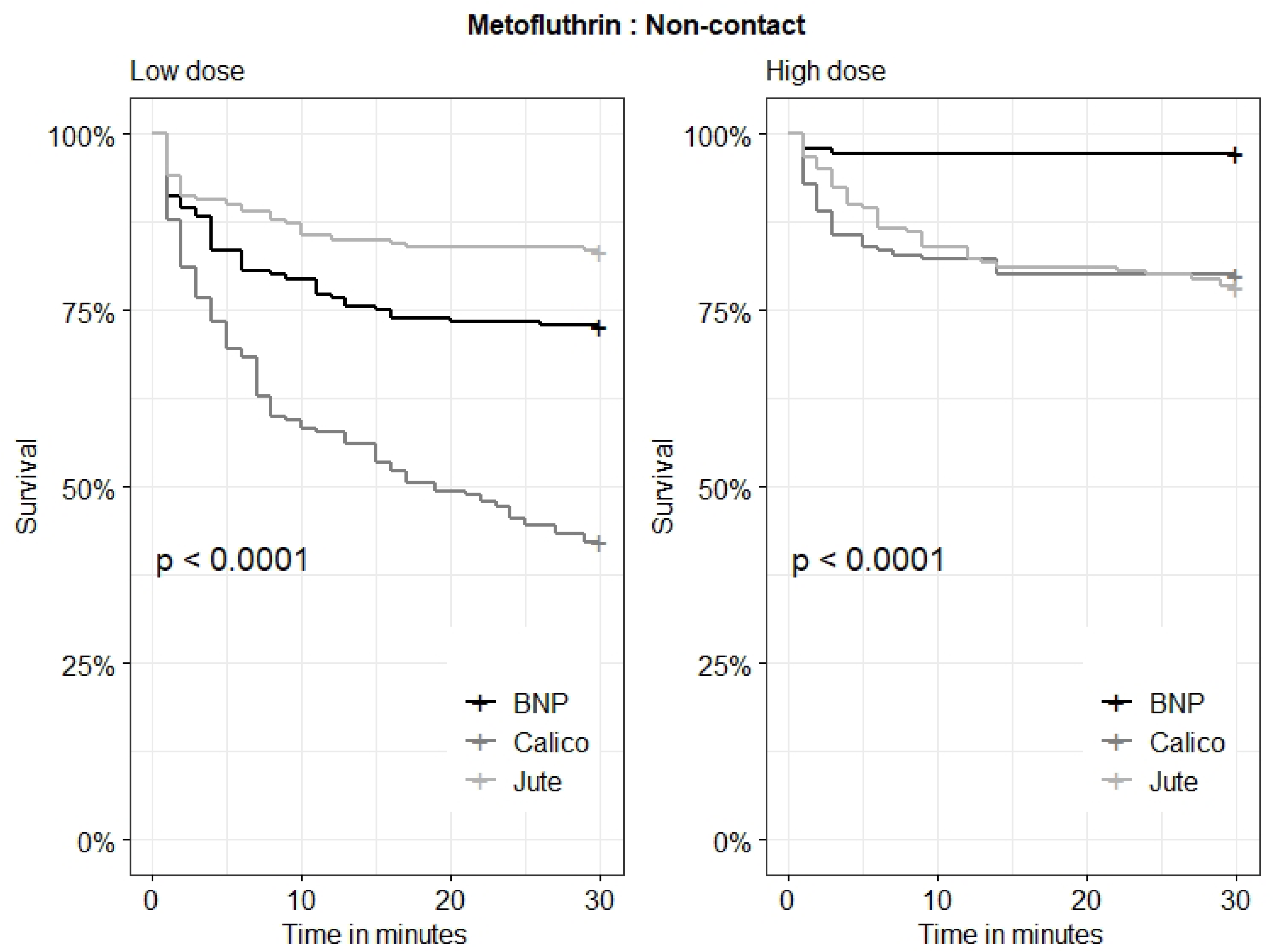
Kaplan–Meier survival curves showing the escape response of *Aedes aegypti* across three metofluthrin-treated fabrics in the non-contact chamber. The curves represent the proportion of mosquitoes remaining unescaped (% survival) over time, with escape treated as the event. Statistical significance was evaluated using the log-rank test, with differences among fabric types considered significant at *p* < 0.05. The actual *p*-value is indicated on the figure.

**Fig 3.**
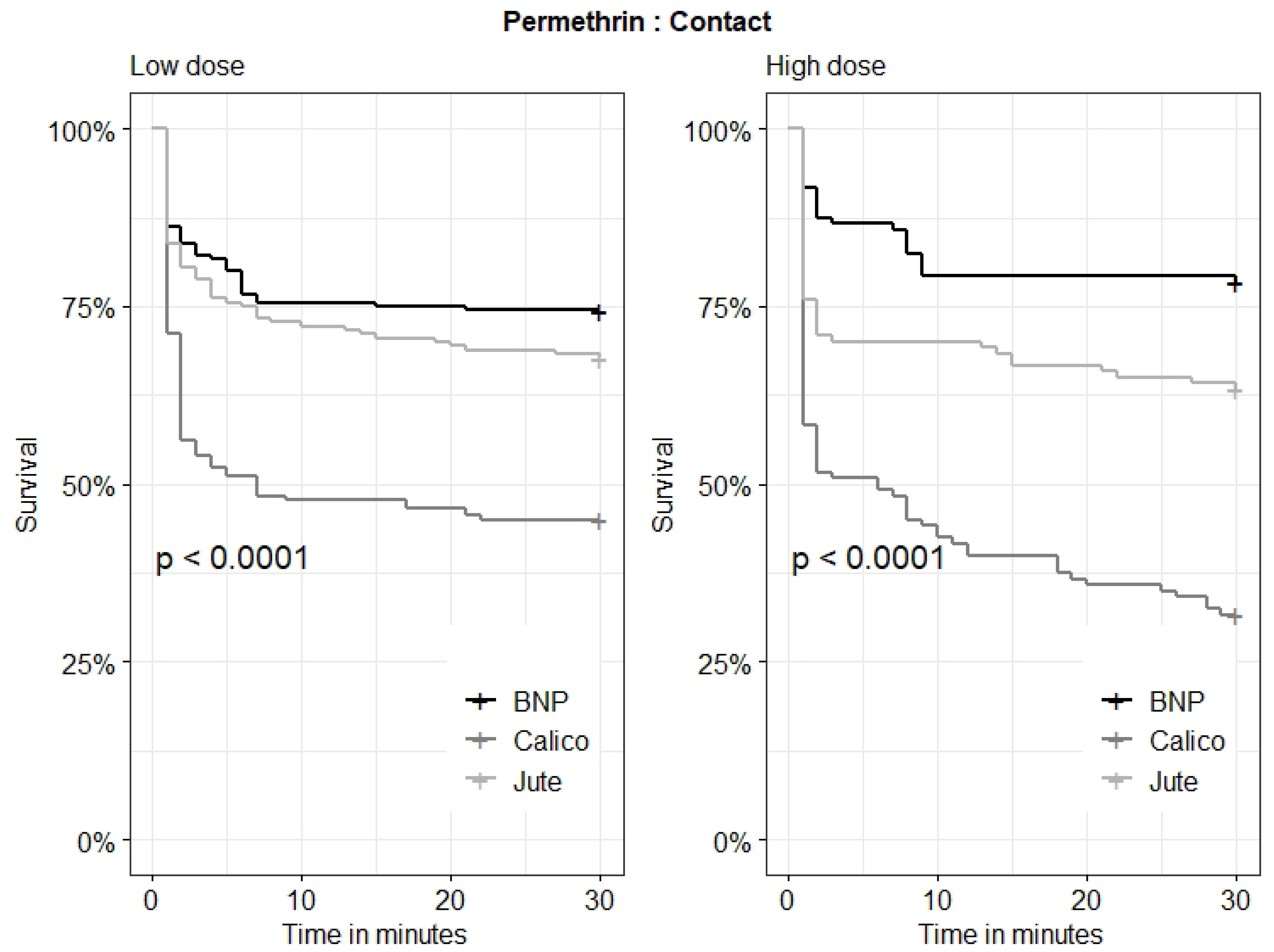
Kaplan-Meier survival curves for escape of *Aedes aegypti* across three permethrin-treated fabrics in contact chamber. The curves represent the proportion of mosquitoes remaining unescaped (% survival) over time, with escape treated as the event. Statistical significance was assessed using the log-rank test. Differences among fabric types were considered significant at ***p < 0.05***. The actual p-value is displayed on the figure.

**Fig 4.**
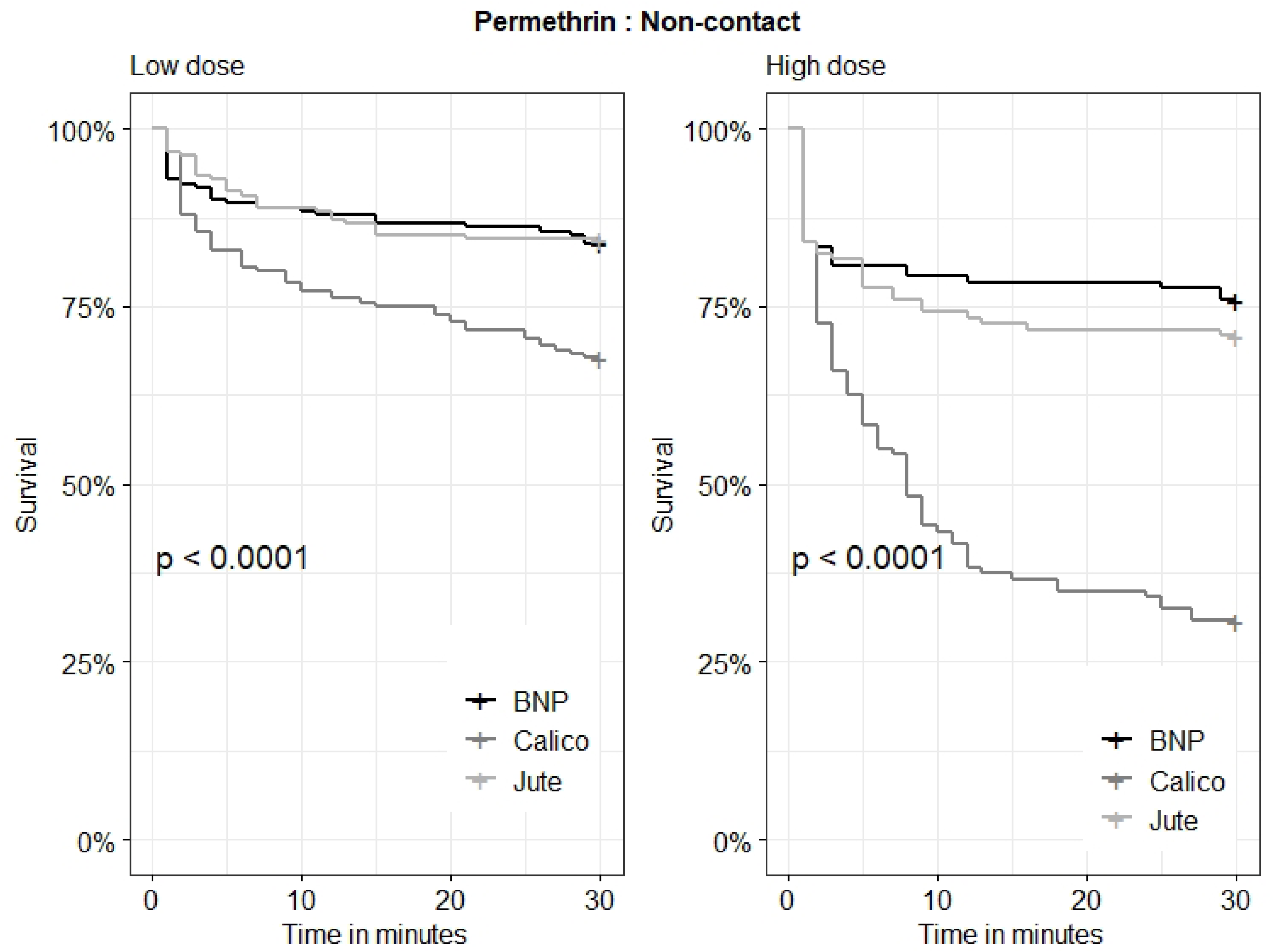
Kaplan-Meier survival curves for escape of *Aedes aegypti* across three permethrin-treated fabrics in non-contact chamber: The curves represent the proportion of mosquitoes remaining unescaped (% survival) over time, with escape treated as the event. Statistical significance was assessed using the log-rank test. Differences among fabric types were considered significant at ***p < 0.05***. The actual p-value is displayed on the figure.

**Fig 5.**
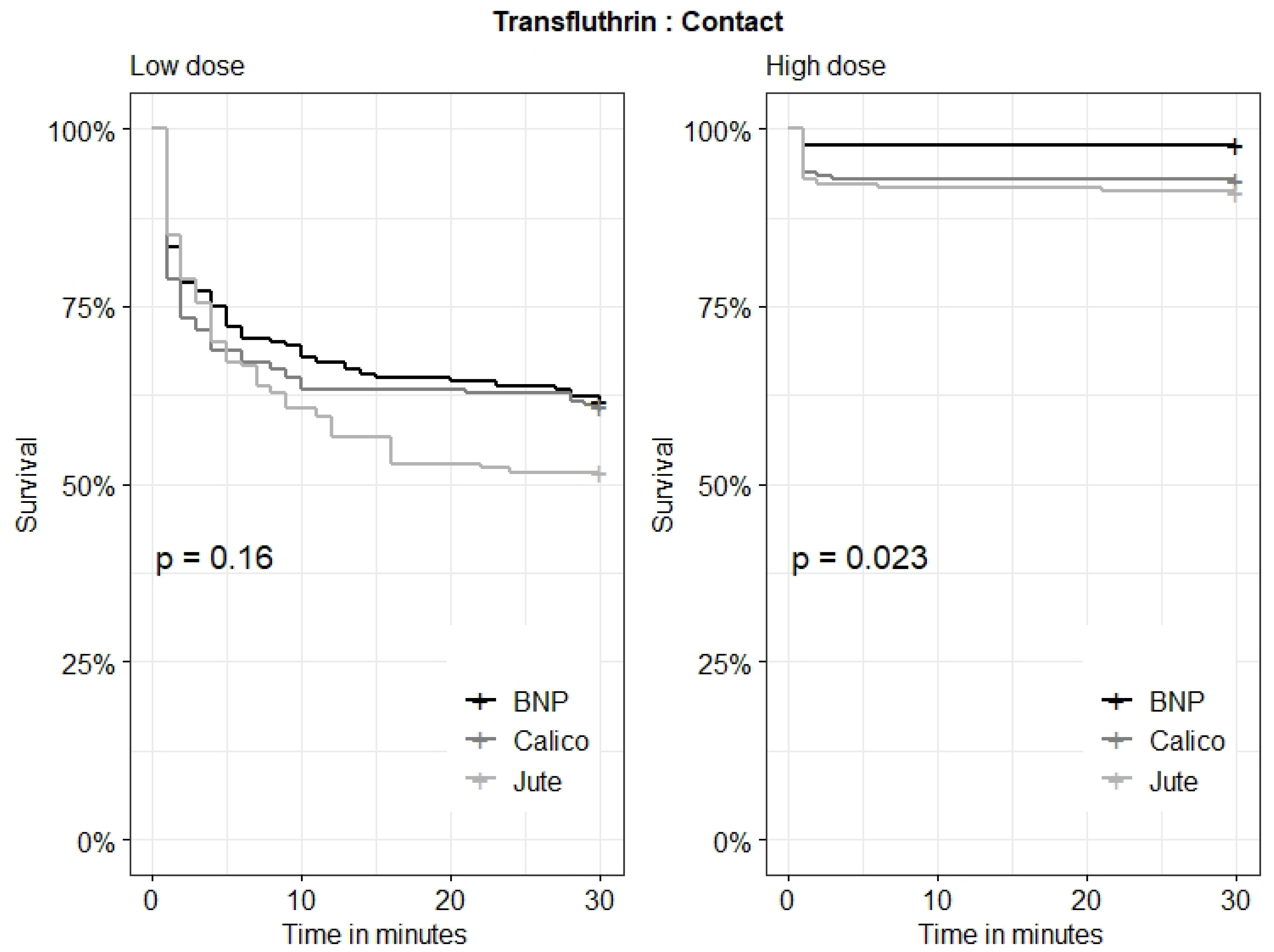
Kaplan-Meier survival curves for escape of *Aedes aegypti* across three transfluthrin-treated fabrics in contact chamber: The curves represent the proportion of mosquitoes remaining unescaped (% survival) over time, with escape treated as the event. Statistical significance was assessed using the log-rank test. Differences among fabric types were considered significant at ***p < 0.05***. The actual p-value is displayed on the figure

**Fig 6.**
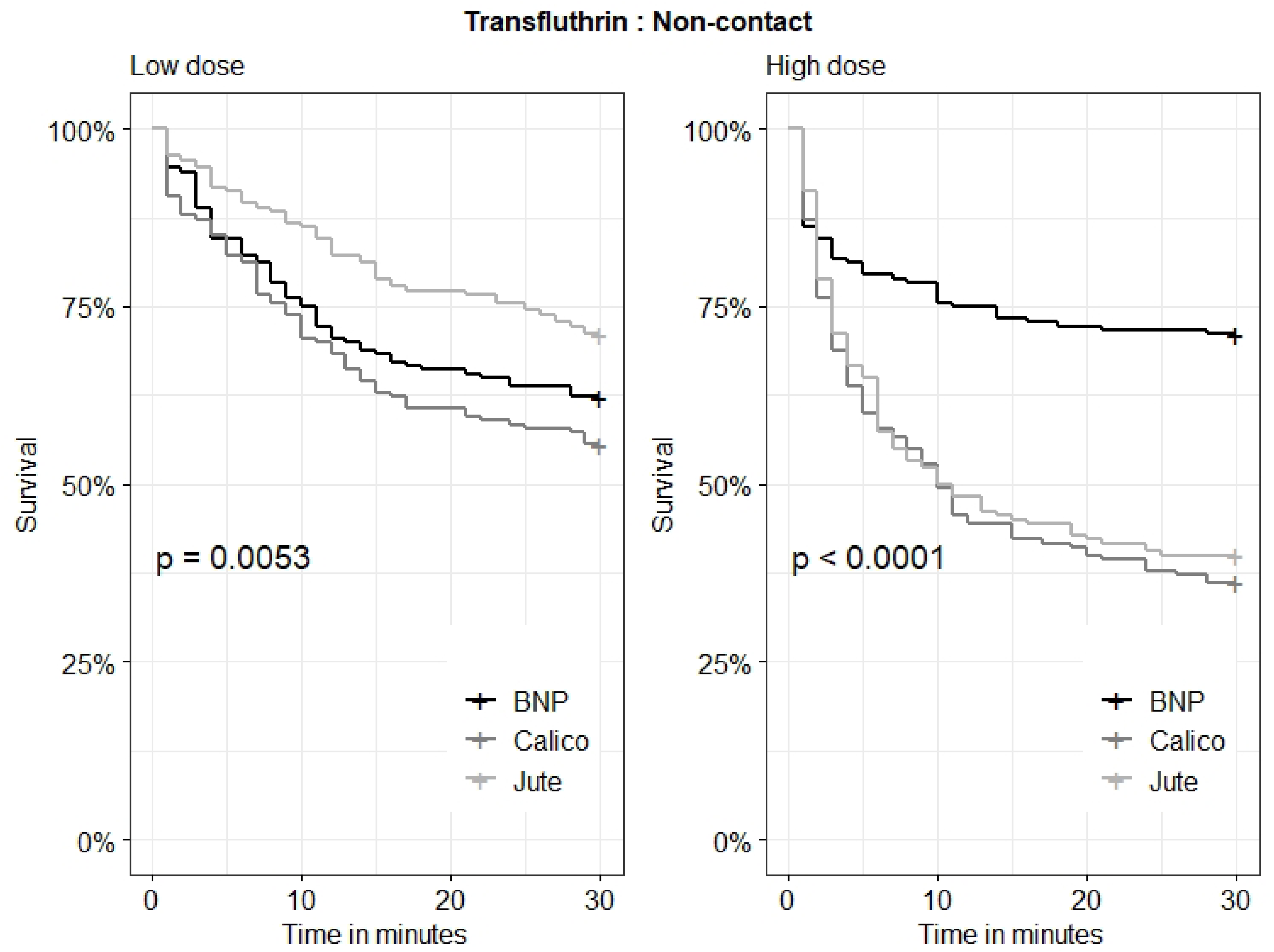
Kaplan-Meier survival curves for escape of *Aedes aegypti* across three transfluthrin-treated fabrics in non-contact chamber: The curves represent the proportion of mosquitoes remaining unescaped (% survival) over time, with escape treated as the event. Statistical significance was assessed using the log-rank test. Differences among fabric types were considered significant at ***p < 0.05***. The actual p-value is displayed on the figure.

Overall, Calico-treated fabrics demonstrated the strongest spatial repellency and irritancy effects against *Ae. aegypti*, while BNP consistently showed the lowest escape rates.

### Toxicity

In GLMM for knockdown and mortality results (Tables 2 and S1, respectively), revealed hat contact exposure was the dominant driver of toxicity. Knockdown was strongly associated with contact trials (OR = 49.17; 95% CI: 40.02–60.41), metofluthrin treatment (OR = 32.27; 95% CI: 22.26–46.79), and high dose level (OR = 9.32; 95% CI: 7.81–11.12). Mortality followed a similar pattern, with elevated odds under contact exposure (OR = 58.88; 95% CI: 47.27–73.34), metofluthrin (OR = 5.26; 95% CI: 3.78–7.30), and high dose (OR = 13.12; 95% CI: 10.96–15.71).

**Table 2.** Estimates of knockdown on *Aedes aegpti* at 24 h post exposure to insecticide-treated fabrics using binomial GLMM.

| Variable |  | Estimate | SE | p-value | OR | LB | UB |
| --- | --- | --- | --- | --- | --- | --- | --- |
| (Intercept) |  | -2.912 | 0.141 | *** | 0.054 | 0.041 | 0.072 |
| Fabric | Cotton | -2.471 | 0.180 | *** | 0.085 | 0.060 | 0.120 |
|  | Jute | -1.091 | 0.165 | *** | 0.336 | 0.243 | 0.464 |
| Test type | Contact | 3.895 | 0.105 | *** | 49.17 | 40.02 | 60.41 |
| Dose category | High | 2.232 | 0.090 | *** | 9.316 | 7.806 | 11.12 |
| Insecticide | Metofluthrin | 3.474 | 0.190 | *** | 32.27 | 22.26 | 46.79 |
|  | Metofluthrin* Cotton | 1.170 | 0.251 | *** | 3.221 | 1.968 | 5.271 |
|  | Metofluthrin * Jute | -1.087 | 0.241 | *** | 0.337 | 0.210 | 0.541 |
|  | Transfluthrin | -0.100 | 0.156 | 0.520 | 1.105 | 0.815 | 1.500 |
|  | Transfluthrin *Cotton | 1.614 | 0.231 | *** | 5.021 | 3.196 | 7.890 |
|  | Transfluthrin * Jute | -0.288 | 0.222 | 0.194 | 0.750 | 0.485 | 1.159 |

SE = Standard error; OR = odds ratio; LB = 95% confidence interval lower bound; UB = 95% confidence interval upper bound
\*Generalized linear mixed model with binomial error distribution and logit link function. Fixed effects include insecticide type, fabric type, test method (Contact vs. Non-contact), and dose category. A random intercept was included for replicate to account for repeated measures.
\*The intercept represents the baseline condition (reference insecticide, fabric, and test type). All odds ratios are relative to these reference levels. OR > 1 indicates increased odds of knockdown; OR < 1 indicates reduced odds.

### Chemical analysis

GC-MS analysis was conducted to assess the retention ability of Jute, Calico and BNP treated with transfluthrin and permethrin during the 1 h and 24 h drying periods. The area composition per sample was calculated for each active ingredient along with the retention times. Transfluthrin concentration was relatively stable in jute fabric (75.7% at 1 h and 76.53% at 24 h) compared to calico and BNP which exhibited large differences between the two time periods. The relative area composition of *cis-* and *trans-* permethrin extracted from the three fabric was generally less variable for the two time periods (Fig 7). A slight variation in the retention times of the analyzed compounds was observed between the two drying periods but the same for the tested fabrics.

**Fig 7.**
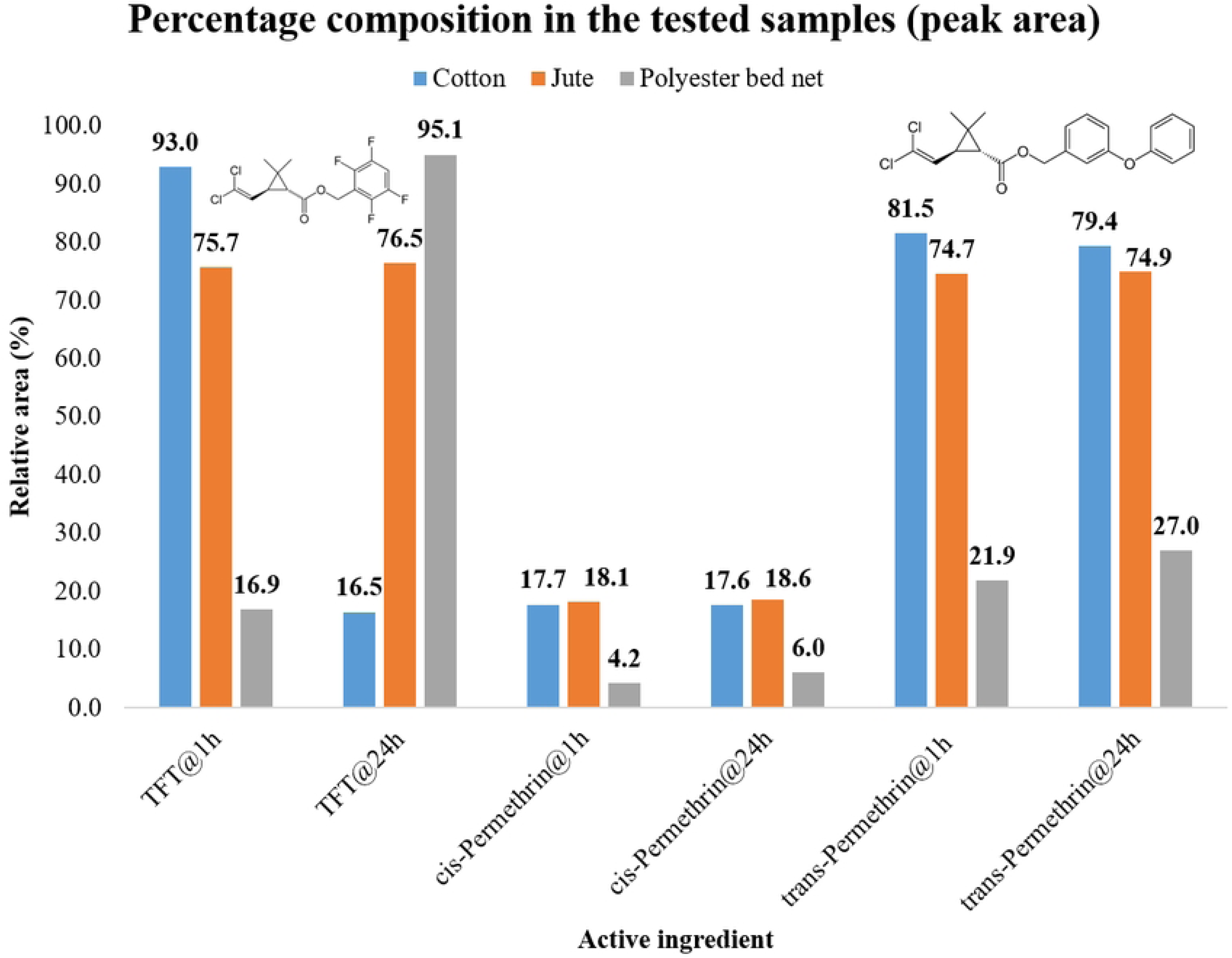
GC–MS analysis of transfluthrin and permethrin retention on jute, calico, and BNP fabrics at 1 hour and 24 hours post-drying. Retention times (min) were as follows: Transfluthrin: 7.86 (1 h), 7.87 (24 h); cis-permethrin - 22.74 (1 h and 24 h); trans-permethrin - 23.08 (1 h), 23.09 (24 h)

## Discussion

Insecticide-treated clothing has long served as a frontline defense against arthropod bites, particularly in military and occupational settings [10, 20, 46]. While permethrin-treated fabrics are well characterized for their dual repellent and insecticidal effects, the role of volatile spatial repellents in treated textiles remains poorly defined. This study is among the first to apply a laboratory-based excito-repellency system to compare the spatial and contact efficacy of three distinct repellent-treated fabrics against *Aedes aegypti*.

By distinguishing between repellency at a distance (spatial repellency) and repellency via physical contact (irritancy) [47], this current study demonstrated that spatial repellency was generally more pronounced than irritancy **(Table 1)**—with the notable exception of permethrin-treated fabrics **(Fig 3)**. This pattern likely reflects the volatile nature of spatial repellents, which activate olfactory-mediated avoidance behaviors prior to contact [5, 24]. In field applications, spatial repellency translates into protective zones that reduce human–vector contact [5], especially during daytime or outdoor activities where ITNs are impractical. This is critical for *Ae. aegypti*, whose diurnal biting behavior limits the efficacy of traditional bed nets [8]. In contrast, irritancy requires direct tarsal engagement with treated surfaces, a response that may be delayed or suppressed by rapid knockdown [31] due to insecticide toxicity.

Toxicity estimates from the present study revealed that contact exposure increased knockdown and mortality by 49.2 and 58.9 times, respectively, compared to non-contact trials. Notably, metofluthrin-treated fabrics produced 32.27 times higher knockdown and 5.26 times greater mortality than permethrin-treated baseline **(Tables 1 and S1)**. These findings are consistent with Ritchie and Devine [48], who reported 80–90% mortality in rooms treated with polyethylene emanators impregnated with 5–10% metofluthrin. Similarly, Kim et al [49] observed high knockdown and mortality using metofluthrin-treated filter papers against *Ae. aegypti*, reinforcing its potent toxicological profile. Excito-repellency assays by Sukkanon et al [31] and Yan et al [16] further support these observations, showing that transfluthrin-treated papers induced high knockdown and reduced escape rates in both *Ae. aegypti* and *Anopheles minimus*. These results suggest that toxicity can mask irritancy where rapid incapacitation limits the opportunity for escape. In our study, spatial repellency was also affected by toxicity at high doses, indicating a complex interplay between dose, volatility, and fabric-mediated release. The exception with permethrin-treated fabrics likely reflects its low volatility and strong contact irritancy, which relies on tactile stimulation rather than airborne dispersion [10]. This “hot foot” effect is well-documented in contact bioassays [50], but its spatial efficacy remains limited unless co-formulated with volatile compounds [51]. These findings underscore the importance of differentiating repellent modes of action and optimizing formulations based on intended use—whether for spatial protection, contact deterrence, or toxicity-driven control.

Calico consistently produced the strongest escape response across trials **(Table 1)**, reinforcing its potential as a high-performance substrate for ITC. This aligns with findings from a previous study [52], where transfluthrin-treated calico repelled *Ae. aegypti* more effectively than polyester or poplin, achieving up to 70% repellency in high-throughput screening assays. Anuar and Yusof [12] reviewed mosquito-repellent textiles and noted that natural fibres like Calico tend to retain repellent agents more effectively than synthetic blends, such as BNP, due to their hydrophilic nature and fibre porosity. The enhanced efficacy of Calico may be attributed to its physical and chemical properties. As a plain-woven, unbleached cotton fabric, Calico offers a porous, hydrophilic surface that facilitates deeper absorption and slower release of active ingredients especially for compounds like transfluthrin and permethrin [53].

Jute demonstrated comparable spatial repellency to Calico when treated with transfluthrin at a high dose **(Fig 6),** irritancy at low dose **(Fig 5)**, suggesting that its coarse, fibrous structure may facilitate effective volatilization of highly active compounds. Unlike Calico, which maintained repellency across insecticides and doses, Jute’s performance was inconsistent— highlighting the importance of fabric–insecticide compatibility. Previous studies have demonstrated that transfluthrin-treated Jute strips reduced mosquito human landing in semi-field trials, but emphasized the need for high-dose formulations to achieve consistent protection [14, 54]. These findings suggest that while Jute may serve as a viable alternative to cotton in spatial repellent applications, its efficacy is contingent on the volatility and concentration of the active ingredient. Operationally, Jute could be prioritized for high-dose transfluthrin formulations in outdoor or semi-enclosed settings [18], whereas Calico offers broader versatility across insecticide classes and exposure modes.

Our findings were reinforced by the GC-MS analysis results **(Fig 7)**, which revealed fabric-specific retention dynamics for transfluthrin and permethrin. Volatile repellent-treated fabrics have been observed to lose efficacy as time passes [55, 56], a factor we hypothesised could have led to the variations in our results after air-drying the fabrics for 24 h before each bioassay. Permethrin concentration was relatively the same across the 24 h period for all the three fabrics. Sullivan et al [56] reported a similar result where they observed unchanged absorption of permethrin on calico (65%)/polyester (35%) fabrics for one week and one month post-treatment. The stable retention in

Calico likely contributed to its superior performance in both irritancy and spatial repellency trials. In contrast, transfluthrin—a highly volatile pyrethroid—exhibited fabric-dependent retention dynamics. Its concentration remained stable on Jute but varied significantly on Calico and BNP between the two drying periods. These fluctuations were inconsistent and did not follow a predictable decay pattern, suggesting that fabric structure and chemical affinity may influence volatilization and bioavailability [13]. Comparable observations were reported by Ogoma et al [55] , who found undetectable airborne transfluthrin at 1 h but variable concentrations at 24 h in rooms treated with jute strips, despite identical treatment protocols.

## Conclusion

Repellent-treated fabrics offer a promising complement to *Ae. aegypti* control strategies due to their affordability and ease of deployment. Present study highlights fabric-dependent differences in repellency and irritancy, with Calico and Jute outperforming BNP across most conditions. While bed net polyester may offer community-level protection due to its disarming potential [28], Calico and Jute are strong candidates for integrated vector management in settings where personal protection is prioritized. Standardizing drying times and conducting further chemical retention studies will be essential for optimizing volatile repellent performance.

**Acknowledgment**

## Data Availability

All relevant data are within the manuscript and its Supporting Information files.

